# Automated Detection of Faciobrachial Dystonic Seizures Related Events in LGI1 Autoimmune Encephalitis Patients with Wearables

**DOI:** 10.1101/2025.03.07.25323205

**Authors:** Jie Cui, Andrea Duque-Lopez, Gabriella Brinkmann, Boney Joseph, Louis Faust, Andrea Stabile, Julianna Ethridge, Gregory Worrell, Divyanshu Dubey, Benjamin Brinkmann

## Abstract

**Objective:** To evaluate the potential of wrist-worn wearable devices to detect and quantify Faciobrachial Dystonic Seizures (FBDS) and related events associated with leucine-rich glioma Inactivated-1 (LGI1)-IgG autoimmune encephalitis (LGI1 AIE).

**Method:** Seven patients and four control subjects were monitored with Empatica E4 wristbands in both hospital and ambulatory environments. The analysis focused on the pre- and post-immunotherapy signals of accelerometry (ACC), electrodermal activity (EDA), heart rate (HR), and blood volume pulse (BVP). A two-stage semi-supervised machine learning approach was developed, utilizing a proprietary algorithm and Support Vector Machine (SVM) classifier to identify FBDS-related events.

**Results:** Significant differences were observed in the characteristics of signals recorded in patients with FBDS compared to controls during sleep periods. LGI1 AIE-associated abnormal events were more frequent, persisted longer, and generated higher ACC amplitude compared to post-immunotherapy and arousal events in the control group. Elevated tonic and phasic EDA were noted in patients, particularly before and after immunotherapy, with a notable decrease in mean and median EDA activity post-treatment, correlating with reduced limbic activation. No significant changes were observed in HR and BVP.

**Significance:** The findings affirm the potential for accurate and automated detection of FBDS and its related events using wearable devices, offering a non-invasive method to quantify seizure burden and treatment efficacy. This approach could minimize the logistical challenges of in-hospital monitoring and provide continuous, decentralized means, improving patient care and clinical decision-making. Future research should focus on expanding the method to daytime monitoring and comparing its effectiveness with in-hospital video-EEG and EMG polygraphy.

**Key points:**

- Assessed the potential of using wearable technology to detect and monitor high-frequency Faciobrachial Dystonic Seizures (FBDS) and related events in anti-LGI1 autoimmune encephalitis.
- Developed a two-stage, automated machine learning algorithm to automatically isolate and classify FBDS-associated events using signals recorded with Empatica E4 wristbands.
- Observed significant differences in the wearable signals of patients with FBDS between pre- and post-immunotherapy, and between patient signals and normal arousal signals of the control group during sleep periods.
- Demonstrated the feasibility of wearable devices to provide objective measures of FBDS-related events, aiding in the quantification of treatment response and influencing clinical decision-making.

## Introduction

Faciobrachial Dystonic Seizures (FBDS) are a common symptom of anti-leucine-rich glioma inactivated-1 (anti-LGI1) autoimmune encephalitis (AIE), a rare form of encephalopathy affecting approximately 14 individuals per 100,000 annually in the United States. These seizures often precede cognitive impairment with confusion and memory loss.^1^

Recording FBDS events is crucial for early recognition and timely intervention to prevent disease progression^2^ and for monitoring the effectiveness of immunotherapy.^3^ Early intervention and accurate quantification of the response to immunotherapy are essential for ensuring effective treatment of autoimmune conditions.^1, 4, 5^ However, in-hospital video-EEG (vEEG) monitoring, the standard approach for other forms of seizures, is expensive and logistically challenging for patients, and thus lasts only a few days, providing limited information on a patient’s clinical state. FBDS are frequent and often not associated with a clear EEG correlate, making their accurate quantification a significant challenge in both patient care and clinical trials.^4, 6^ Additionally, the accuracy of patient-reported seizure diaries is often questionable. Studies indicate that patients frequently under-report their seizures, with estimates suggesting that fewer than half of seizures are reported.^7-11^ This issue is particularly concerning for FBDS events due to their high frequency.^12^ Furthermore, tracking seizures during sleep poses significant difficulties.^13^

Wearable devices have proven successful in noninvasively monitoring focal to bilateral tonic-clonic seizures.^14, 15^ However, their efficacy in detecting other seizure types, including FBDS, remains unkown.^14, 16^ This study aims to evaluate the potential of using wristworn devices to detect the characteristic limb movements and physiological changes related to FBDS. This could provide valuable insights into disease severity and treatment effectiveness. To date, no study has successfully used wearable sensors to distinguish FBDS from normal movements and nocturnal sleep arousals. The use of non-invasive wearable devices to detect and quantify FBDS could provide a vital measure of seizure burden and treatment efficacy for this challenging condition.^17^

## Method

The Mayo Clinic Institutional Review Board (22-006702) approved this study. We recruited seven patients (P1-P7) with LGI1 autoimmune encephalitis (AIE), all of whom provided written informed consent before participating. ^1^TABLE 1 summarizes the demographic and clinical information of the cohorts. Participants were asked to wear a wristworn physiological monitoring device (Empatica E4, Empatica Inc., Milan Italy) that collected 3-axis accelerometry (ACC), electrodermal activity (EDA), photoplethysmography (PPG)-derived blood volume pulse (BVP), and skin temperature (TEMP). Heart rate (HR) and inter-beat-interval (IBI) were derived from BVP.^15^ The device was worn on the limb most affected by FBDS events, when bilateral asynchronous FBDS were present.

For data analysis (TABLE 2), the sleep periods of eight hours for patients P1 and P2 were identified around the start of their immunotherapy (P1-a and P2-a); the sleep periods of four patients (P3-5 and P7) were identified posterior to immunotherapy (range: 9 – 406 days). The sleep period of P6 was identified six days prior to the start of immunotherapy. All these periods were chosen at the earliest time when E4 monitoring was available. P1 and P2 had an additional sleep period analyzed (P1-b and P2-b), approximately five months and one month, respectively, following their first sleep period collection using the same E4 device. We chose overnight periods for the analyses to minimize the confounds of activity-related movement during the day and due to the poor quality of self-reporting events during sleep.^13^ More detailed descriptions of patients P1 and P2 are provided below, as two sleep periods were analyzed in their course of treatment, which can reveal more clinically relevant information.

**TABLE 2.** Timing of the recordings in relation to symptom onset, immunotherapy, recording duration, detected event count, and event frequency in seven patients with anti-LGI1 autoimmune encephalitis and four control subjects.

| Subject | From onset to therapy (d) | From therapy to monitoring (d) | Empatica E4 recording for analysis (a [b]) |  |  |  |
| --- | --- | --- | --- | --- | --- | --- |
|  |  |  | Since therapy (d) | Recording time (h) | Event number | Event frequency (/h) |
| P1 | 35 | 0 | 0 [153] | 8 [8] | 109 [66] | 13.63 [8.25] |
| P2 | 81 | 0 | 0 [33] | 8 [7] | 199 [71] | 24.88 [10.14] |
| P3 | 0 | 406 | 406 | 7 | 39 | 5.57 |
| P4 | 85 | 77 | 77 | 9 | 22 | 2.44 |
| P5 | 445 | 262 | 262 | 5 | 10 | 2.00 |
| P6 | 1271 | -6 | -6 | 7 | 11 | 1.57 |
| P7 | 36 | 9 | 9 | 8 | 10 | 1.25 |
| 4 Controls | -- | -- | -- | 29 | 100 | 3.45 |

Column titles: **From onset to therapy**, time from the onset of diagnosed FBDS symptoms to the start of immunotherapy; **From therapy to monitoring**, time from the start of immunotherapy to the start of Empatica E4 monitoring; **Empatica E4 recording for analysis (a [b])**, metrics of analyzed E4 recording number **a** and number **b** (shown in bracket if available); **Since therapy**, time elapsed since the start of immunotherapy; **Recording time**, total recording time of recording **a** [recording **b**]; **Event number**, total number of detected events in recording **a** [recording **b**]; **Event frequency**, average number of events per hour in recording **a** [recording **b**]. Abbreviations: **FBDS**, faciobrachial dystonic seizures; **d**, days; **/h**, number of events per hour. Note: \*E4 monitoring started 6 days before immunotherapy. “--” indicates data are not available.

**Patient P1** is an ^2^age-year-old male experiencing high-frequency right-sided FBDS associated with a brief period of slurred speech. He also had minor cognitive deficits and mood changes. Brain MRI was unremarkable, and LGI1-IgG were positive in serum. After diagnosis, event monitoring with Empatica E4 was performed (see above). Approximately one month after symptoms onset, he received a course of high-dose IV methylprednisolone (1g/day for 5 days) followed by oral prednisone taper (from 80mg/day to discontinuation over 4.5 months), in addition to oral lacosamide (100mg BID). Due to the persistence of FBDS, rituximab was also administered to achieve seizure freedom (TABLE 1).

**Patient P2** is an age-year-old male who developed episodes of sudden contraction of the right face and arm, often followed by the contraction of the same segments contralaterally, consistent with bilateral “a bascule” FBDS. The motor phenomena were often followed by a period of confusion, lasting a few seconds. Initially, these episodes occurred 6-10 times per day and became even more frequent (every 20 minutes) after less than two months. Oral levetiracetam (250mg BID) was ineffective. Apart from minor cognitive and personality changes, he also experienced episodes of orthostatic hypotension with falls. Antibody testing found LGI1-IgG positivity in serum. CSF analysis showed only an increase in proteins (80mg/dL, normal range 0-35), without pleocytosis. Brain MRI demonstrated a subtle T2/FLAIR hyperintensity in the mesial temporal lobes. Seizures were monitored using an Empatica E4 device on the left wrist. Approximately 2.5 months from clinical onset, immunotherapy was administered. Despite the use of first-line (IV methylprednisolone 1000mg/day for 5 days, then 1000mg/week for 8 weeks, 5 cycles of plasmapheresis)h, plasmapheresis) and second-line agents (rituximab), the patient continued to have FBDS, albeit with reduced frequency (1-2 FBDS per day, TABLE 1).Four control subjects (CS), including one normal volunteer (Control 1) and three patients with epilepsy (Control 2-4) were selected from an existing cohort for comparison. Control 2 was monitored in both an epilepsy monitoring unit at Mayo Clinic and home, while Control 3 and 4 were monitored at home with seizure confirmation provided by an implanted NeuroPace RNS EEG recording device.^17^ Control data for these patients were analyzed from periods where they were confirmed to have had no seizures or other events, and wearable data were aggregated for statistical analysis.

Four control subjects (CS), including one normal volunteer (Control 1) and three patients with epilepsy (Control 2-4) were selected from an existing cohort for comparison. Control 2 was monitored in both an epilepsy monitoring unit at Mayo Clinic and home, while Control 3 and 4 were monitored at home with seizure confirmation provided by an implanted NeuroPace RNS EEG recording device.^18^ Control data for these patients were analyzed from periods where they were confirmed to have had no seizures or other events, and wearable data were aggregated for statistical analysis.

Given the common lack of EEG signals modifications correlated with FBDS events, inhospital EMU video-EEG monitoring may be ineffective in FBDS quantification. To address this limitation, a two-stage approach was developed to identify potential events of interest. An in-house feature-based algorithm (FIGURE 1) was first used to identify events from the pre-treatment data of P1 and P2 (P1-a and P2a, identified around the immunotherapy start TABLE 2), who had the highest frequency of event occurrence. While it is not possible to be certain that individual events were FBDS or FBDS-related events, they represent abnormal high-frequency motor-involved events only observed in LGI1 AIE, who reported and were observed having active FBDS. The same algorithm was employed to identify non-FBDS-related events, presumably representing normal sleep arousal, within the control cohort. A machine learning algorithm was then trained on the identified labeled events to separate them, which was then used to classify events in the post-treatment data of P1 and P2 (P1-b and P2-b), as well as the datasets of P3 through P7.

**FIGURE 1.**
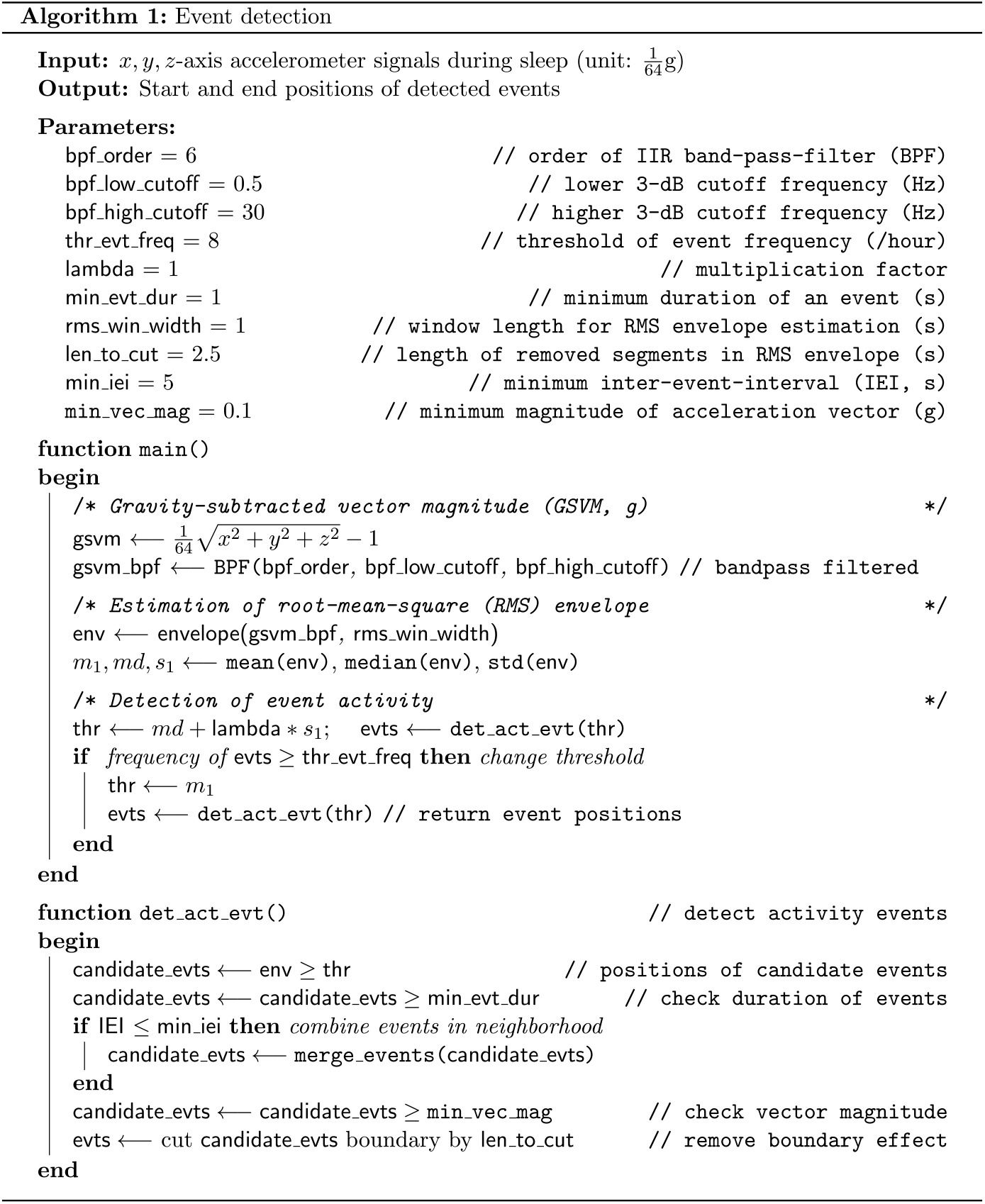
ACC event detection algorithm. The algorithm processes the *x, y*, and *z*-axis signals from an Empatica E4 accelerometer recorded during sleep periods. The output is the timestamps of the detected start and end times. First, the algorithm calculates the GSVM from the original accelerometer signals and uses an IIR filter to remove baseline drifting and high frequency noise. It then estimates the RMS envelope and its mean, median, and standard deviation from the filtered signals. Finally, these metrics are input into the action event detection algorithm det_act_evt(), which identifies candidate events if the RMS envelope exceeds a threshold for consecutive periods longer than a predefined minimum duration. Adjacent candidate events with short gaps are merged, and those with magnitude above a minimum amplitude are considered detected events. The start and end timestamps of these vents are reported after removing boundary effects. Abbreviation: **IIR**, infinitive impulse response; **BPF**, band-pass filter; **RMS**, root-mean-square; **IEI**, inter-event-interval; **GSVM**, gravity-subtracted vector magnitude; **STD**, standard deviation.

The initial stage of the algorithm identified events of interest using the Gravity-Subtracted Vector Magnitude (GSVM) of ACC signal in units of *g*, calculated as 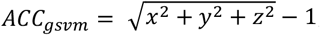, where *x, y* and *z* are the three components of the E4 accelerometry signals sampled at 32 Hz. A bandpass filter with cutoff frequencies at 0.5 and 30 Hz was applied to the ACC signal to remove baseline drift and artifacts. The Root-Mean-Square (RMS) energy envelope was then estimated from the filtered ACC using a sliding window of length 5 seconds and thresholded at one standard deviation (SD) above the mean to identify candidate events. If the event frequency exceeded 8 events per hour, a level at which we observed that continuous arousals were observed to be fragmented, the threshold was then reduced to the mean and the candidate events were re-calculated. Candidate events shorter than 1 second and with low amplitude (< 0.1*g*) were removed. Detected events with inter-event-intervals (IEI) of less than 5 seconds were aggregated. Finally, half the length of moving window (2.5 seconds) was trimmed from the onset and end points of candidate events to avoid boundary effects. The algorithm is summarized in FIGURE 1, with examples of the detected events shown in FIGURE 2.

**FIGURE 2.**
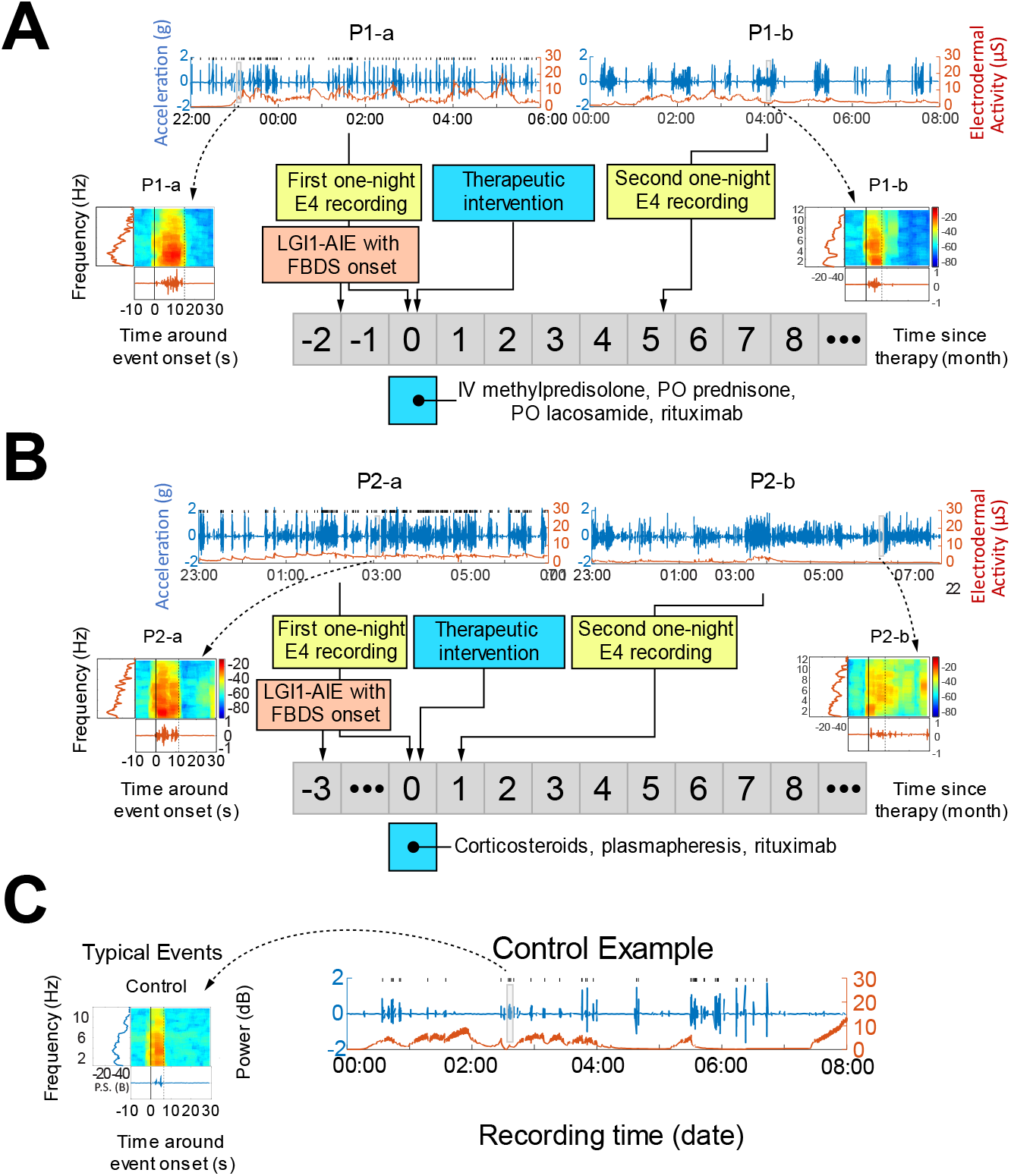
Clinical timelines, ACC, EDA, and identified events for two LGI1 patients and one control example. **(A)** Displays the ACC (blue curve, left axis) and EDA (red curve, right axis) of two one-night E4 recordings of patient P1 (P1-a and P1-b) taken at various times compared to the start of immunotherapy. P1-a was recorded at the beginning of immunotherapy (0^th^ month), and P1-b was recorded approximately 5 months posterior to the therapy (5^th^ month). The array of numbers shows the time elapsed since the start of the therapy in months (negative number indicate time before the therapy start, 0 indicates the month of immunotherapy, and positive numbers indicate time after the therapy). See TABLE 2 for more details. The blue square below shows the approximate duration of immunotherapy and the prescribed medication (listed on the right side the box). The inlets on both sides show the time-frequency representations of the two typical ACC events shown in P1-a and P1-b (gray rectangular boxes), respectively. We define a typical event as the one whose ACC magnitude and duration match the medians of all detected events. (**B**) Shows a similar graph for patient P2. Note that the two E4 recordings (P2-a and P2-b) were separated by approximately one month. This patient was not seizure-free during the E4 monitoring period (see TABLE 1). (**C**) Displays an example of ACC and EDA signals and a time-frequency representation of the typical event of a control subject. Abbreviations: **ACC**, accelerometry; **EDA**, electrodermal activity.

In the second stage, upon identifying the event envelope, we calculated the duration, rising time, and falling time of the detected events, as well as BVP, EDA, and TEMP characteristics during events. The duration was defined as the time interval between the onset and the end of an event. The rising time was determined as the elapsed time for the envelope to reach half of the envelope maximum from onset, and the falling time was calculated from the envelope maximum to half the maximum. The multitaper method^19^ was used for frequency and time-frequency analysis of the event dynamics.^20^ The EDA signal was high-pass filtered at 0.05 Hz to isolate the phasic component^21^, which was then subtracted from the original EDA signal to obtain the tonic activity component of the signal.

Using the identified abnormal LGI1-associated and normal events, we trained a Support Vector Machine (SVM) classifier based on three features of each event: GSVM, event duration, and the Skin Conductance Level (SCL) of EDA signals. A Gaussian kernel was employed. We evaluated several kernel scales, and a scale of 0.43 produced the best results. The classifier was trained using a 5-fold crossvalidation, with the misclassified rate serving as the loss function (MATLAB fitcsvm function).

## Results

The study identified an average sleep duration of 6.72±0.96 (mean ± SD) hours for the control cohort and 7.44±1.13 hours for the patients (TABLE 2). For the SVM model training, 100 non-FBDS and 308 FBDS were identified. The LGI1 patients recruited for the study exhibited varying degrees of disease severity (TABLE 1). Four of them (P3-5 and P7) had undergone various degrees of immunotherapy treatment prior to E4 monitoring due to the clinical urgency to initiate immunotherapy upon diagnosis.

The identified FBDS-related events typically exhibited a longer duration and higher energy compared to control events (FIGURE 1, FIGURE 3B, FIGURE 4A, B). The classifier achieved an accuracy of 0.88 and an AUC of 0.82 during training (FIGURE 3A). In total, 229 events were identified as potential LGI1-associated abnormal events from 51 hours of recordings of P1-a, P2-a, and P3 through P7 (FIGURE 3). It was evident that the dynamic range of the control events was much smaller than the potential abnormal events identified by the algorithm when comparing control and pre-treatment data of P1 and P2 (FIGURE 4B, C & D). The distribution of abnormal events showed greater dispersion in the feature space than the controls.

**FIGURE 3.**
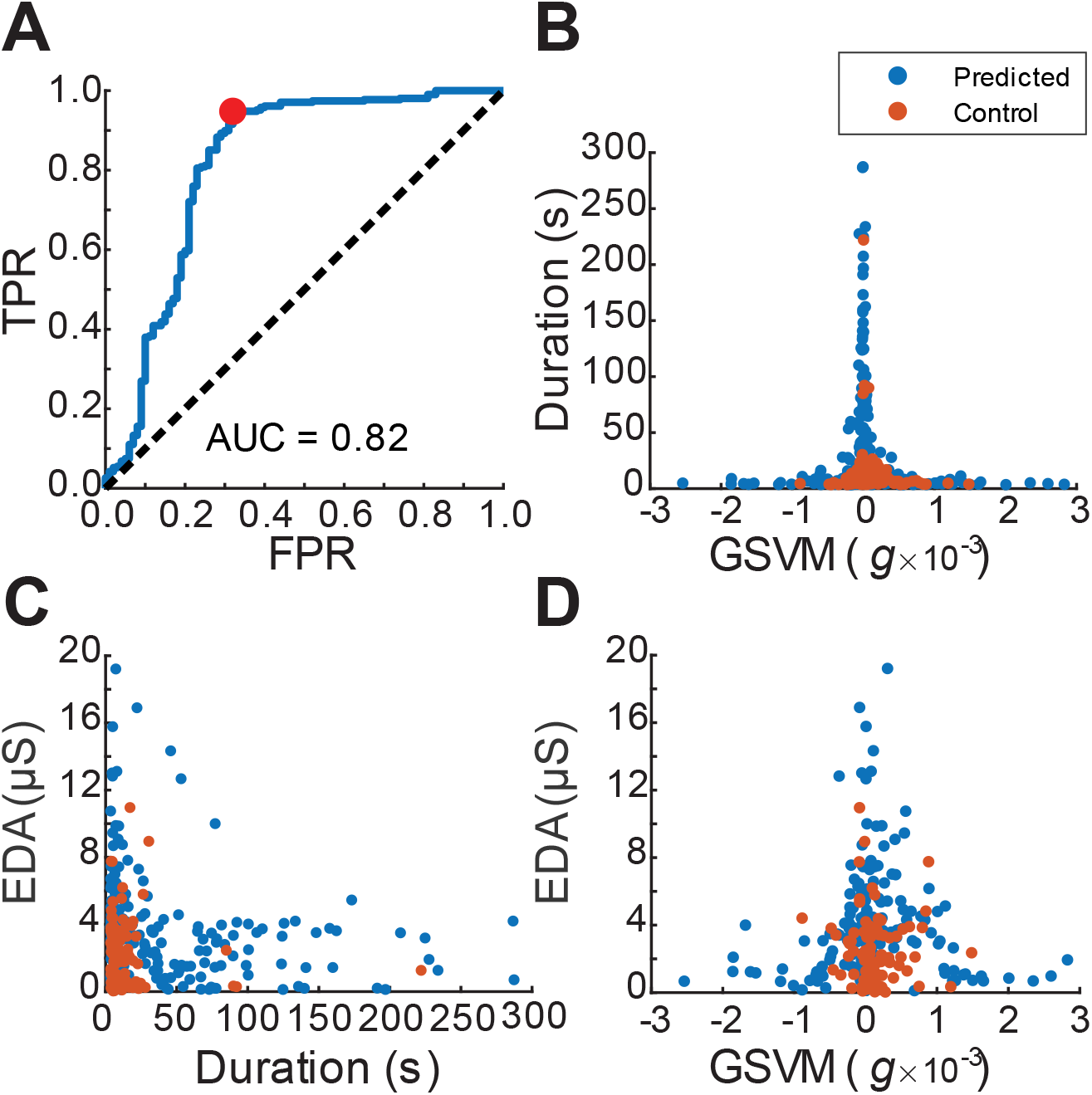
SVM-based prediction of FBDS events with Gaussian kernel. **(A)** Displays the ROC curve of the SVM classifier (refer to text for details). The red dot indicates the operating point. The figure displays scatter plots of the detected events in the control group and predicted events in the sub-feature space of **(B)** GSVM and duration, **(C)** duration and EDA-SCL levels, and **(D)** GSVM and EDA levels. Abbreviation: **FPR**, false positive rate; **TPR**, true positive rate; **GSVM**, gravity-subtracted vector magnitude; **EDA**, electrodermal activity; **SCL**, skin conductance level.

**FIGURE 4.**
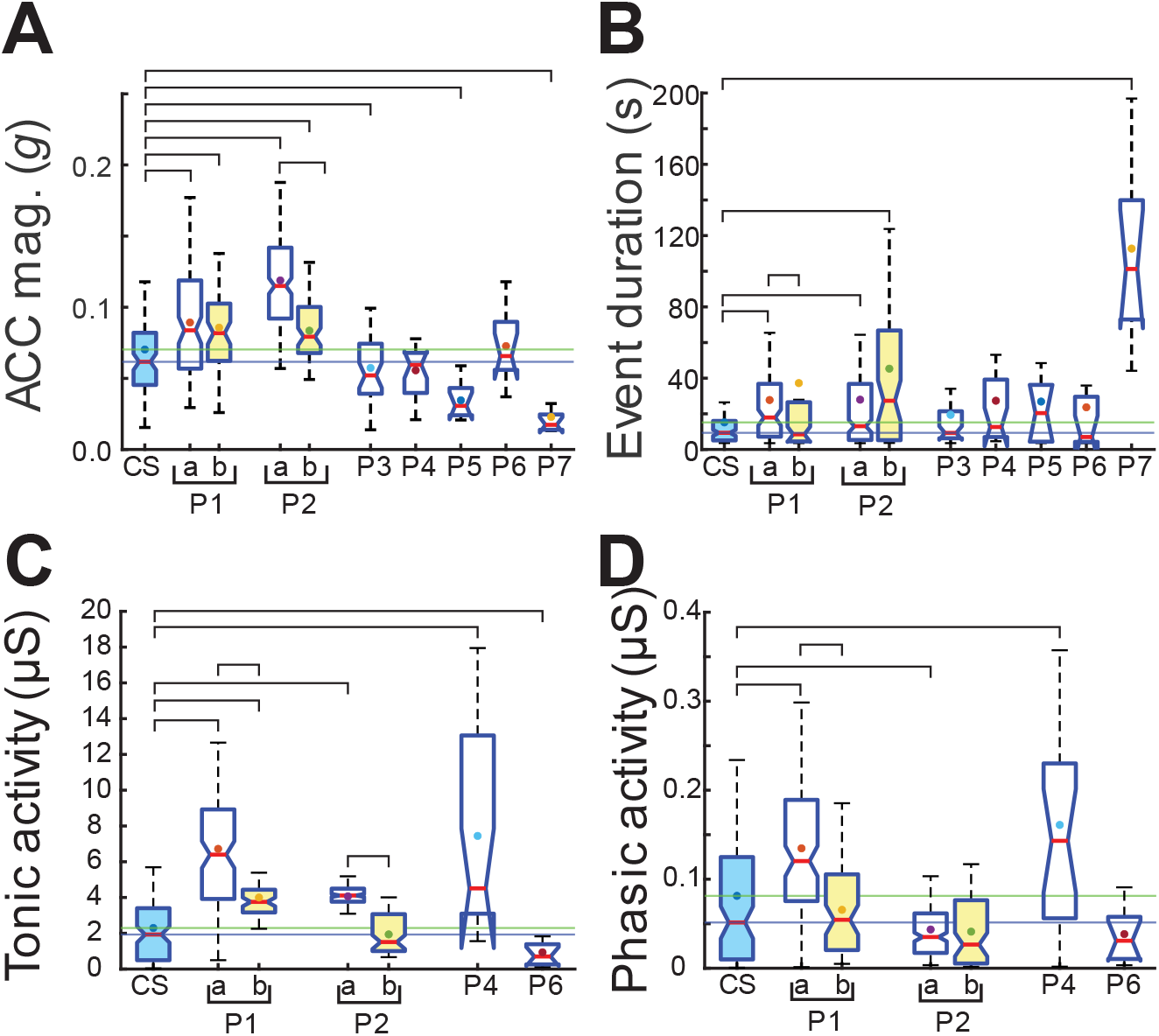
Boxplots of detected and predicted event actigraphy. Event **(A)** ACC magnitude, **(B)** duration, **(C)** tonic activities, and **(D)** phasic activities are shown for the controls (CS, blue boxplot) and the seven patients (P1-P7). Note that P3, P5 and P7 lack reliable EDA measurements during the predicted events. Overhead horizontal bars denote statistically significant differences between the medians of the corresponding pairs (Wilcoxon rank sum test, all *p* < 0.05). Abbreviation: **ACC**, accelerometer; **mag**., magnitude; **P1-a**, first one-night E4 recording of P1; **P1-b**, second one-night E4 recording of P1 (yellow boxplot); **P2-a**, first one-night E4 recording of P2; **P2-b**, second one-night E4 recording of P2 (yellow boxplot) We also found significant differences in the median event duration (FIGURE 4B) between the control group and the FBDS-related events in the pre-treatment phase for both patients (P1-a and P2-a vs. Control, 27.76±32.89 s and 27.92±34.64 s vs. 15.17±25.69 s, *p* < 0.05). However, in the post-treatment phase, a significant difference in the median was only observed in P2-b, not in P1-b. Moreover, we found a significant difference between pre- and post-treatment in P1. Among patients P3-P7, only the median of P7 shows significant difference.

We found that the estimated frequency of actigraphy events (events/hour, /h) in the pre-treatment sleep period of P1 (13.63±4.24 /h) and P2 (24.88±13.52 /h) were significantly higher than that of control group (3.45±3.00 /h) (two-sample t-test, *p* < 0.001). Notably, the event frequency post-treatment (P1-b and P2-b) was significantly lower than pre-treatment (P1-a and P2-a) in both patients (TABLE 2), reflecting the effect of immunotherapy. However, the event frequencies of P3-P7 did not show a significant difference from the controls.

In the actigraphy analysis of detected events, we observed significant differences in ACC magnitude between the FBDS event and controls (FIGURE 4A). Interestingly, the ACC magnitude for both P1-b and P2-b are smaller than their pre-treatment counterparts (P1-a and P2-a) and were closer to the control group after treatment. Although the ACC magnitudes of P3, P5 and P7 were also different from controls, they were smaller, not larger as in P1 and P2, indicating an increase in variance in FBDS-related event features.

For both tonic and phasic activity of EDA signals (FIGURE 4C, D), we found a significant difference in means between pre-treatment and control (P1-a vs. Control, *p* < 0.001; P2-a vs. Control, *p* < 0.001), and between post-treatment and control (P1-b vs. Control, *p* < 0.001; P2-b vs. Control, *p* < 0.001) in both patients. The medians of the tonic and phasic activity after treatment were lower than those before treatment and were closer to the control group in both patients. For patients without the second one-night sleep analysis (P3-P7), the median of P4 was significantly higher, but P6 significantly lower, than the controls, which might suggest increased variance in FBDS-related events. We could not analyze reliable tonic/phasic activities of EDA levels for P3, P5 and P7.

No significant differences were observed between the LGI1 AIE and controls in HR, BVP or TEMP during the detected events.

## Discussion

This study demonstrates significant difference in the characteristics of signals recorded by a wearable device in patients with anti-LGI1 AIE and FBDS compared to control subjects during sleep periods. These findings support the potential for more accurate and automated detection of FBDS and related events, as well as for quantifying FBDS burden and treatment efficacy in patients. LGI1-associated abnormal events were more frequent, persisted longer, and generated higher ACC amplitudes than arousal events in the control group and after immunotherapy. These events also exhibited significantly elevated tonic and phasic EDA compared to control and post-immunotherapy events, particularly evident in P1 and P2 with analyzed pre- and post-treatment recordings. Notably, both mean and median EDA activity decreased after treatment in both patients, correlating with the anticipated reduction in limbic activation after treatment.

Although all patients were diagnosed with ‘LGI1-AIE with FBDS’ (TABLE 1), two patients (P5 and P6) did not exhibit clinically observed FBDS. Their medical history suggested a possible occurrence of FBDS prior to clinical observation, but no clinical FBDS was documented, leaving it uncertain whether FBDS occurred during E4 monitoring. This uncertainty might explain the relatively low frequency of detected FBDS-related events in these two patients (TABLE 2).

While no significant changes were observed in HR or BVP for these patients, these signals may still be useful to monitor, as heart rate variability has been implicated in autoimmune inflammatory encephalitis,^22^ and considerable heterogeneity may exist between patients. Interestingly, while the first LGI1 patient (P1) reported seizure freedom following the course of immunotherapy, he also reported a possible new onset of rapid eye movement (REM) sleep behavior disorder, which might explain why the nocturnal event count remained high post-treatment.

The stereotypical movements involved in FBDS, often without a clear electrographic discharge on scalp EEG, make these paroxysmal motor phenomena well-suited for monitoring with wearable devices. These devices may be particularly beneficial for monitoring during nighttime hours when patients and caregivers may be asleep and unable to keep a record of events. Our proof-of-concept analysis benefitted from the limited movements during overnight hours. However, some caution is needed when assessing patients with bilateral asynchronous FBDS (as P2), as the device may not detect some episodes. Future development would expand to assessing the method’s ability to identify FBDS events during daytime hours and to compare its effectiveness with in-hospital vEEG monitoring and with EMG polygraphy. The wearable device’s capability to measure EDA, HR, and TEMP in addition to ACC may facilitate the detection of other semiologies associated with LGI1-IgG seropositivity, such as pilomotor seizures. A possible accurate quantification of seizure events is critical to guide therapy and measure response in autoimmune epilepsy, a considerable challenge in most AIE trials. ^5^

The outcome of this study demonstrates the possibility to quantify seizure burden and cognitive symptoms through an entirely remote procedure, minimizing the logistical challenges of in-hospital monitoring. This procedure could be incorporated into clinical practice to assist in titrating the dose and duration of immunotherapy in autoimmune encephalitis. With the capabilities of modern wearable devices, it is possible to mail devices to patients and configure them remotely, acquiring usable data without requiring the patient to physically return to the clinic.

Continuous, non-invasive, and minimally burdensome decentralized monitoring over extended periods, is a considerable advantage in conditions like this, as it reduces the burden on patients who may find it difficult to travel, while offering accurate and objective data to guide clinical decision-making “outside the clinic”.^16, 23^

A limitation of this study is the absence of gold-standard labels for FBDS events, which poses a challenge to traditional supervised machine learning approaches. These events occur frequently and may be missed in self-reported diaries. Several machine learning approaches within the framework of weakly labeled learning^24^ may offer potential solutions to this problem, which are currently under investigation. Traditionally, assessing the overall seizure burden and efficacy of treatment is clinically more important than analyzing individual events for patient management. However, the technology of wearable devices offers new potentials for precise timing and quantification of disease status, the impact of which on clinical practice warrants further investigation.

## Conclusion

This study has demonstrated the potential of using a wrist-worn wearable device to automatically detect FBDS-related events in patients with LGI1 autoimmune encephalitis (AIE). The detection relied on the characteristics of the accelerometric and EDA signals. The identified events were notably different from similar events observed in controls and post-immunotherapy in the patients. Our investigation has affirmed the feasibility of quantifying the response of LGI1 AIE patients with FBDS to immunotherapy, thus providing objective evidence to guide treatment strategies and influence clinical decisions. However, additional research is necessary to improve the accuracy of FBDS event detection.

## Author contributions

**Jie Cui**: Funding acquisition, conceptualization, data analysis, and composition of the manuscript. **Andrea Duque-Lopez**: Data collection and manuscript revision. **Gabriella Brinkmann**: Data collection and manuscript revision. **Boney Joseph**: Conceptualization and data collection. **Louis Faust**: Manuscript revision. **Andrea Stabile**: Data analysis and manuscript revision. **Julianna Ethridge**: Study coordinator and data collection. **Gregory Worrell**: Funding acquisition. **Divyanshu Dubey**: Funding acquisition, conceptualization, and manuscript revision. **Benjamin Brinkmann**: Funding acquisition, conceptualization, and composition of the manuscript.

## Acknowledgement

The authors acknowledge Sherry Klingerman, CCRP, Yvonne Larson, and Mona Nasseri, PhD, for technical and regulatory support. The authors dedicate this manuscript to the memory of Nicole McClure, who helped with coordination of this study.

## Data availability statement

Limited data are available upon reasonable request to the authors.

## Footnotes

1 TABLE 1 of demographic information is not available in medRxiv preprint due to medRxiv polidy.

2 Precise age number is not available in medRxiv preprint due to medRxiv policy.

